# Implementation and evaluation of synchronous chat in general practice - a qualitative interview study

**DOI:** 10.1101/2025.03.27.25322992

**Authors:** Elin. J. Schelling, Reinier C.A. van Linschoten

## Abstract

**Introduction:** General practices in the Netherlands are experiencing high workload due to rising healthcare demand and staff shortages, particularly in rural areas. Digital healthcare innovations are being explored as solutions to alleviate these challenges. However, evidence regarding their effectiveness is limited.

**Methods:** This qualitative study explored the implementation and effects of a synchronous chat application, Uw Zorg Chat, in general practices. Semi-structured, in-depth interviews were conducted with healthcare workers involved in a pilot program in Leeuwarden. Interviews were transcribed, pseudonymized, and analyzed thematically.

**Results:** The application’s design and triage functionality were well-received, but issues with collaboration between practices and the Uw Zorg Chat team created challenges, mainly due to changes in workflow. Practices less prepared for change, with missing team alignment and a lack of feedback structures, viewed these changes negatively. Although some practices noted a slight reduction in workload, it was unclear whether this was attributable to the app. In contrast, other participants perceived an increased workload due to the need to monitor and correct the work performed by Uw Zorg Chat. Patient adoption of the app was lower than anticipated, limiting the observable effects.

**Conclusion:** Definitive conclusions about Uw Zorg Chat’s effectiveness in reducing workload cannot be drawn, as the limited patient adoption resulted in minimal impact. However, the study highlights the importance of organizational readiness for change and the need to establish strong communication channels during implementation to support successful integration.

## Introduction

In the Netherlands, general practitioners (GPs) play a crucial role in managing patient care as the first point of contact within the healthcare system. They handle 93.3% of healthcare problems independently, addressing a wide range of issues without the need for referral to secondary care (1). When necessary, they also act as gatekeepers, referring patients to specialized care to ensure efficient use of healthcare resources. However in recent years the pressure on general practice has significantly increased, leading to challenges regarding accessibility and quality of care. Studies show that 82% of GPs and 76% of doctors assistants (DAs) experience high levels of work pressure, with pressure likely to rise further in the future (2). This pressure harms patient care, patient-provider communication and patient safety (3, 4).

In addition to the increasing workload, more than half of Dutch general practices are anticipating staff shortages (2). These shortages are particularly concerning in suburban and rural areas, where attracting sufficient physicians has already proven challenging (5). This issue is exacerbated by a declining interest among young GPs in becoming practice owners, which has traditionally been an important aspect of general practice in these regions (6). Many newly graduated GPs opt to work as locums or in salaried positions, often remaining in urban areas where they completed their training (7). Consequently, the continuity of care, especially in rural areas, is in jeopardy, as practices are forced to close when older GPs retire and no successors are found.

Together with these capacity challenges comes an increasing demand for healthcare, driven in part by an aging population and the associated rise in chronic conditions and multimorbidity (8). Projections indicate that the demand for GP services will substantially increase in the coming years, while the supply of GPs and support staff will not be able to meet this demand (9). This creates a concerning situation where the balance between healthcare demand and supply is increasingly at risk of becoming unbalanced, particularly in regions where care is already vulnerable.

To address capacity challenges in general practice, the Dutch government is increasingly turning to digital healthcare solutions. Digital healthcare encompasses a wide range of tools, including electronic consultations, telemonitoring, chat services, digital triage, and electronic data exchange. These innovations aim to improve care delivery while alleviating the workload of healthcare providers (10, 11)

A prominent concept within digital healthcare is hybrid care, which combines digital tools with traditional face-to-face consultations. For example, electronic consultations allow patients to send messages to their GP, which are addressed asynchronously within a few days. However, evidence suggests that asynchronous communication may not significantly reduce workload, as inefficiencies arise from delays between interactions (12-15).

Given these limitations, there is a growing interest in synchronous digital communication, such as live chat. This approach enables real-time interaction and allows for quicker triage and resolution of patient inquiries. Although several providers in the Netherlands—such as Arene, Medicoo, and some health insurers—offer synchronous chat options, no studies have yet assessed their impact on GP workload.

This study seeks to investigate the implementation of a synchronous chat application in general practice and its potential to reduce workload.

## Methods

### Qualitative approach and research paradigm

This qualitative study used semi-structured interviews and thematic analysis to explore healthcare workers’ experiences with implementing synchronous chat consultations in general practice to reduce workload. A phenomenological approach was chosen to gain an in-depth understanding of personal experiences, perceived barriers, and potential recommendations to improve the implementation process of a synchronous chat application.

Our research questions were:

- What are barriers and facilitators in the implementation process of a synchronous chat consultation in general practice?
- What effects are noticed after implementing a synchronous chat consultation?

### Researcher characteristics and reflexivity

The researcher, a medical student at Erasmus Medical Center, conducted this study for her master’s thesis. Prior to the study, she worked as a doctor’s assistant, answering chat consultations for a pilot program of a synchronous chat application. This gave her valuable insight into the system, helping her ask focused questions and interpret findings. However, this experience could have introduced bias by influencing her objectivity. To minimize this, she remained aware of potential influences, had reflective discussions with her supervisor, and ensured the analysis stayed grounded in the data. She had no prior clinical or supervisory relationships with participants, ensuring her neutrality as a researcher.

### Context

This study examines synchronous chat consultations in general practice. A pilot program was implemented from July to October 2024 in four general practices in Leeuwarden using a synchronous chat application called Uw Zorg Chat. These practices faced medical staff shortages, particularly GPs, during the summer months. To address these shortages, a chat-based application was introduced, allowing patients to contact healthcare workers via live chat with free text. The staff managing the chat were located in Rotterdam and were not affiliated with the local practices or familiar with the patients. Upon receiving a message, a doctor’s assistant triaged the inquiry and could provide advice, refer the chat to an online doctor, or schedule an appointment at the patient’s own practice. Appointments made through the chat system were given a different color in the scheduling system from those made directly at the practice. All conversations were documented in the practices’ systems. Contacts requiring a prescription or referral were managed by the GPs at the participating practices, while all other inquiries were handled by Uw Zorg Chat. Patients were informed about the possibility to use the chat application through automated phone messages and flyers in the participating practices. In October 2024 the pilot ended, but one of the participating practices decided to continue with the use of Uw Zorg Chat, while the others did not. This study was conducted after the pilot phase, and participants were asked to reflect on their experiences with this synchronous chat application.

### Sampling Strategy

The study population consisted of staff (e.g. GPs, doctors assistants [DAs], practice managers) of the four participating general practices. Employees were excluded if they commenced working after the start of the pilot. All staff were invited to participate in the study via e-mail and WhatsApp by the practice managers. After accepting the invitation, we assessed whether the participant met the inclusion criteria. We aimed to include all eligible employees of the general practices that participated in the pilot to ensure sufficient depth and diversity in the data. When no new topics emerged during an interview, two additional interviews were conducted to confirm data saturation. If new topics arose in these subsequent interviews, the process was repeated until no further new themes emerged. Data saturation was assessed separately for the practice that continued and those that stopped to capture any differences in their experiences.

### Ethical issues pertaining to human subjects

The research question originated from Uw Zorg Chat, where the researcher was formerly employed. The research was conducted independently, without financial or other compensatory arrangements. Additionally, the researcher had not been worked for Uw Zorg Chat since the start of the study. A protocol was submitted to the Medical Ethics Committee (METC) of Erasmus Medical Center and deemed exempt from the Medical Research Involving Human Subjects Act. Informed consent, both written and oral, was obtained from all participants prior to the interviews. Participants received an informed consent letter after accepting the invitation to participate in the study, and oral consent was confirmed before the interviews commenced. Participant confidentiality was maintained through data pseudonymization and the secure storage of interview recordings and transcripts.

### Data Collection Methods

Semi-structured in-depth interviews were conducted in November and December 2024. A prompting and probing interview technique was employed, using open-ended questions and encouraging participants to elaborate on their views. The interviews were guided by a topic guide developed beforehand (see Supplementary File 1), based on the Nonadoption, Abandonment, Scale-up, Spread, and Sustainability (NASSS) Framework (16). This guide was designed to be flexible, allowing adjustments as new themes emerged during the discussions. The initial aim was to conduct all interviews face-to-face, recorded via MS Teams, in consultation rooms at the participating practices. However, when face-to-face interviews were not feasible, online MS Teams meetings were conducted as an alternative. Each interview lasted approximately 45 to 60 minutes. Following data collection, the interviews were transcribed verbatim and subsequently pseudonymized by assigning participant codes. Baseline data were collected from all participants, including age, gender, role (e.g., GP, DA practice manager), years of experience at the participating general practice, and prior experience with digital health modalities (e.g., e-consultation, email, telephone, video, telemonitoring, digital triage; yes or no).

### Data Processing and Analysis

The interviews were recorded and transcribed verbatim, with each participant assigned a unique code number to ensure data pseudonymization. The transcripts were then imported into MAXQDA for analysis. The process began with open coding, focusing on phrases relevant to the research questions. A coding tree was developed in MAXQDA, based on the topic guide, the results of open coding and the NASSS Framework (16). This coding framework was reviewed and refined in collaboration within the research team. Axial coding followed, offering a clearer structure of (sub)themes.

### Techniques to enhance trustworthiness

To ensure reliability and accuracy, the first author developed an initial coding tree based on six interviews. Subsequently, a second researcher validated the codes by independently coding two additional interviews, after which the consistency in codes was assessed. Through discussion and refinement, a final coding tree was established. Any need for additional codes was addressed through further discussion between the two researchers.

## Results

### Characteristics of participants

The study included 14 participants of the four participating general practices of whom 10 (71.4%) were female, with a mean age of 51.7 years (Table 1). Among the participants, three were GPs and practice owners of the participating practices, one of them continued the use of Uw Zorg Chat, and two stopped. Three were practice managers, one continued the use of Uw Zorg Chat, two stopped. Five were DAs, three continued the use of Uw Zorg Chat, two stopped. Two were practice assistants specializing in somatic care, one continued, one stopped. One was an advanced nurse practitioner, working in the practice that continued the use of Uw Zorg Chat. The mean duration of work experience in the participating practices was 11.7 years (see Table 1). All participants had experience with digital healthcare modalities, including, email and e-consultations. One practice had previously conducted a hybrid healthcare pilot, which meant that five participants had prior experience with this form of digital healthcare. Additionally, one participant reported experience with telemedicine in a previous role.

**Table 1:** Characteristics of participants.

| **N=14** | **N (%) / years (SD)** |
| --- | --- |
| Female | 10 (71.4%) |
| Mean age | 51.7 (13.7) |
| Mean work experience at practice | 11.7 (8.1) |
| *Experience digital health care* |  |
| Usual (e-mail, e-consultation) | 14 (100%) |
| Hybrid care pilot | 5 (35.8%) |
| Telemedicine | 1 (7.1%) |
| Participants discontinuing use of Uw Zorg Chat after end of pilot | 8 (57.1%) |

### Main findings

The main findings of this study were structured into categories aligned with the research question: the implementation process and the effects. Based on the NASSS Framework (16), a coding tree was developed for the implementation process, themes covered the technology, the value proposition, the adopter system, the organization and the wider context. This resulted in questions about the application Uw Zorg Chat, the collaboration between Uw Zorg Chat and the organization (the general practices), the structures within the general practices and the wider context. Additional questions were asked about the effects, including positive, negative or neutral outcomes. These findings were divided in themes and subthemes (Figure 1). A total overview of codes with definitions and examples can be found in Supplementary File 2.

**Figure 1:**
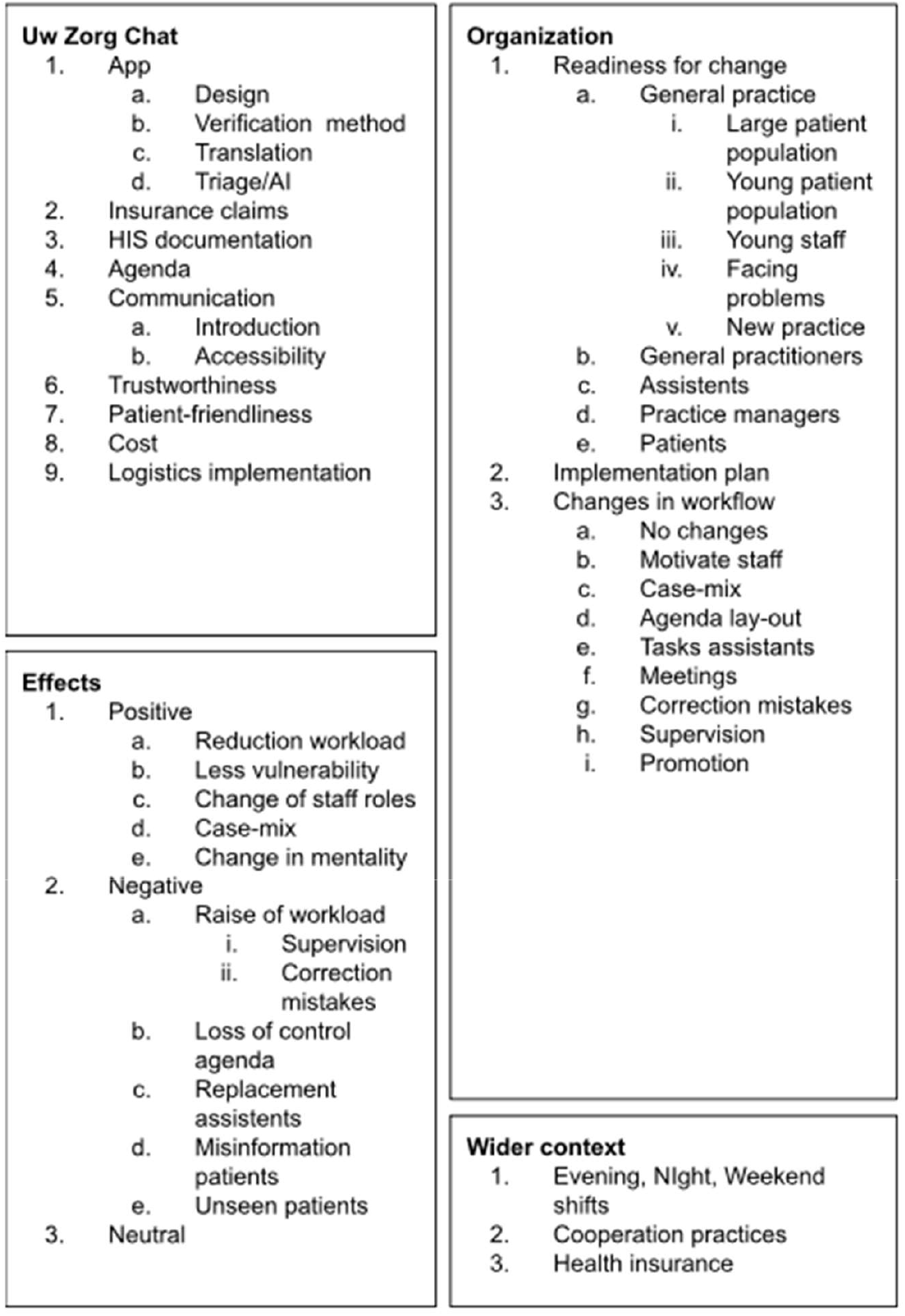
Themes and subthemes coding tree.

### Uw Zorg Chat

This topic includes the application itself, the organization handling the queries, as well as the cooperation and communication between the staff of Uw Zorg Chat and the staff of the participating GP’s practices.

#### The application

The design of the app was generally well-received, particularly the way patients were triaged. One practice manager commented: *“Well, I am in favor of AI, and it was backed by Thuisarts and NHG guidelines.”* (P1, PM) Thuisarts and the NHG guidelines are the officially recognized Dutch clinical practice standards followed by GPs and DAs in patient care. Furthermore, the translation feature was appreciated for improving accessibility for patients.

A frequently mentioned barrier was the account verification process, which relies on iDIN: a Dutch digital identification and authentication service which uses the banking infrastructure. Users found this method challenging to trust, because of the need of a small payment (€0.01). Moreover, this process was not feasible for specific patient groups, such as individuals under financial guardianship, those with outdated phones, or those with foreign bank accounts.

#### Collaboration

The integration of Uw Zorg Chat into the systems of participating practices received mixed feedback. The practice that continued using Uw Zorg Chat reported positive experiences with personnel of Uw Zorg Chat scheduling consultations, documenting contacts in the electronic health record (EHR), and processing insurance claims. They also noted that issues were quickly resolved. The practice owner of the continuing practice mentioned: *“Especially in the beginning, everything that was new—for us, but also for you—was immediately addressed. And also the billing. Like, oh, I see this and that. How are you used to doing it, and how are we used to doing it? And it was corrected. And in the last weeks, it didn’t happen anymore, at least not for this practice—it was just well organized.”* (H1, GP)

Conversely, practices that chose to not continue after the pilot expressed dissatisfaction with how feedback was handled: *“But I found the handling of my complaint—well, I had already mentioned several times in the beginning that I thought billing wasn’t going well. I didn’t think they addressed it adequately. The same issues came up again later.”* (H2, GP)

All DAs in the practices that did not continue highlighted issues with scheduling, notation, and declaration: *“For example, it sometimes happened that patients were scheduled in slots normally reserved for home visits. And often, information was missing, like with that lab form—you had to arrange something, but you couldn’t because the information wasn’t there. Or if we had to make a referral, it wasn’t clear what it was about or where the patient wanted to be referred.”* (D3, DA)

DAs from the continuing practices acknowledged the mistakes but did not view them as significant inconveniences and were open to providing feedback to Uw Zorg Chat. One DA commented: “*Well, we didn’t have any comments, but we did have some questions or unclear things. A few times, we got some administrative things back. That’s not a big deal in itself, but then the documentation of contact was just a bit too short or insufficient. Then we didn’t know what we were supposed to do with it.”* (D1, DA)

Practices that chose not to continue with Uw Zorg Chat operatedas a healthcare center in a single location, comprising four GP practices of which three practices participated in the pilot. The continuing practice is an independent, individual GP practice. One assistant observed: *“Not everything is the same across practices. Everyone works differently. It’s all solo, right? I think if you’re going to implement this in these practices, everyone needs to work uniformly. In the practice that continued, they all work uniformly. We have a very different setup than the other GPs. Everyone works differently. We have different times for home visits, different consultation hours. It’s difficult for someone working for such a service to deal with so many different setups.”* (D4, DA)

Participants identified several facilitators for better collaboration. Many participants expressed a desire for an introduction to the Uw Zorg Chat team. They suggested activities such as a group event or visitation of Uw Zorg Chat workspace. Additionally, they recommended a clearer introduction to the app, for example, through an explanatory video. Participants believed that these measures would help build greater trust in the product and enhance job satisfaction.

To resolve scheduling and feedback issues, participants recommended improving communication between the practices and Uw Zorg Chat. One participant explained: “*Look, I understand it’s difficult with scheduling, but I would have liked it if they could just call and say: do I see correctly that I can plan this here, or am I misunderstanding it? We are very easy to reach for consultation. I think I sometimes missed that: how are we going to approach this, or what’s the best way to handle it?”* (D3, DA)

#### Trust

Most of participants expressed trust in the app and the team behind it: *“Yes, exactly, as I said before, we had more trust here. Right away. And the assistants too, because we didn’t have to do much. Not even triage.”* (D1, DA) However, some participants struggled with losing control over the scheduling process. A practice manager mentioned about her DAs: “*In the beginning they would still call patients to see if the information was correct, as it was entered into the system. I talked about it with one of the assistants. I said, you know, if you don’t think the other side is competent enough, we have to stop using the app, you have to let that go and then she let it go.”* (P3, PM)

#### Patient-friendliness

Patients shared many positive responses, and doctors who tested the app as “patients” themselves were satisfied with both the advice provided and the quality of interactions. The app was particularly valuable for patients seeking answers to minor questions. Accessibility was also highly praised by participants. *“The patients are very satisfied. And what I also notice is that people who are a bit insecure or have minor issues— they tend to ask questions that you might think, ‘You could have asked your mother or grandmother,’ but they say they wouldn’t do that. For those people, this app works beautifully. That experience of having a doctor in your pocket works really well with this app.”* (H1, GP)

#### Costs

The practices that chose to discontinue the pilot primarily cited high costs as the reason. They believed the expenses outweighed the benefits and would have preferred hiring an additional employee at the same cost, as they could handle multiple tasks. Participating practices with smaller patient populations reported that the financial burden was unsustainable for smaller practices. In contrast, the practice continuing with Uw Zorg Chat had a patient population twice the size of the other practices and considered the costs acceptable. Additionally, poor billing practices exacerbated the feeling that the app was a financial drain: *“And there was supposed to be reimbursement, at least for the consult fee, right? If people contacted the chat, it was billed in the EHR. That should cover the difference. But later, we found that we had around 150 consultations, and for 50 of those, it wasn’t even billed. So financially, it was a big problem.”* (H2, GP)

### Organization

#### Readiness for change

##### Practices in general

Several participants from practices that discontinued the pilot questioned whether their practice was well-suited for implementing Uw Zorg Chat. These practices often already had established accessibility solutions, such as online patient portals to get insight into medical data and send e-consultations, catered to small patient populations, and did not face challenges with staff shortages or workload pressure. Additionally, some participants noted that their patient base consisted largely of older individuals, who were thought to be less likely to benefit from an application like Uw Zorg Chat.

##### General practitioners

While GPs were willing to participate and had clear expectations, they were not always prepared to invest effort in the implementation process. Practices that discontinued the pilot were often unwilling to adapt their workflows, such as closing phone lines or actively promoting the app, while these two methods were addressed as the main facilitators to get patients to use the app. *“There were interim evaluations where they said: you need to focus more on A, more on B—things like raising awareness and instructing patients. But that just didn’t happen.”* (P3, PM) This resistance was partly due to a desire to maintain patient service and personal contact: *“As a GP, I think it’s important my patients can reach me directly and not rely on a chat service. My patient population includes older people who are very digitally skilled and younger ones who aren’t, and vice versa. Closing phone lines takes away some accessibility and personal connection. General practice is about personal relationships, where patients value speaking with someone who knows them and their situation.”* (H3, GP).

As previously mentioned, three of participating general practices were in the same building. In the same building a fourth general practice was located. This practice did not want to participate in the pilot, due to the work attitude of the other three practice owners: *“We had done some projects before, and because the four of us were supposed to co-operate, we found it tricky to partner up with the other three. It had failed before. We knew they couldn’t pull it off, because they don’t put in enough effort”* (P2, PM)

The practice that continued with Uw Zorg Chat was led by a forward-thinking practitioner who was open to innovation and willing to take risks. This attitude was mentioned by the staff: *“During the holiday, it was quite calm, so people could easily come in. The practice owner once said, you should let the phone ring, then we’ll force them to use the app.”* (P1, PM)

##### Assistants

DAs of continuing practices were positive about the introduction of Uw Zorg Chat. Many expressed curiosity about the system’s functionality and its potential to improve workflow. They identified benefits such as a change in work: “*In the worst-case scenario, I would only be seeing patients. Well, I wouldn’t mind that at all; I actually enjoy doing that. Of course, I’d miss the phone a little because it’s often fun, but seeing patients is what I enjoy most. So if the phone work completely disappears, personally, I wouldn’t mind.”* (D2, DA). They could see the added value of an application like this: *“Do you think the practice needed an app like this?* (Int.) *I always thought so. It was a very busy practice”* (D2, DA)

The practice continuing with Uw Zorg Chat highlighted that being a large practice facilitated the implementation process. In larger practices, changes can be addressed collaboratively with all DAs, making transitions more manageable. However, implementation was complicated by a prior pilot conducted, which had been perceived negatively. This previous experience made it more challenging to approach a new pilot with confidence, especially given the uncertainty regarding the project’s long-term viability: *“If the pilot ends after three months and we don’t continue with it, and I’ve been recommending it to everyone, it’s like I’ve dangled a carrot they never get. That feels a bit fake.”* (H1, GP). Although they also mentioned positive differences: “*I personally noticed very little of it, actually. And I think that’s because you completely relieve us. The previous practice we worked with still contacted us regularly, they wrote to the patients that they had to call us again. This is actually much more present in the background, so that we don’t actually notice it.”* (D2, DA) .

Of the discontinuing practices, a practice manager noted that some, mostly older, DAs were content with the existing processes and preferred to remain focused on telephone tasks, showing less enthusiasm for change. Additionally, some DAs expressed concerns about the potential redundancy of their roles, fearing that digital tools could replace them, leading to uncertainty about their future: *“Some assistants are worried. Like, ‘Am I still needed now?’ or ‘What if I’m no longer relevant?’ It doesn’t seem to matter if they’re younger or older—it’s just that the unknown feels unappealing.”* (P1, PM)

Some DAs would have preferred more involvement in the decision-making process: *“It all happened quite suddenly, like one day we were just doing this. Later, we had a meeting where we could share our thoughts on what we found important. Still, I sometimes felt there was a lack of communication with us.”* (D3, DA) However, in the practice continuing the pilot, this was less of an issue, as a preparatory team meeting had been conducted to discuss the app’s introduction.

##### Patients

Patients generally appeared to respond positively to change according to the healthcare providers: *“Especially younger people and those who are digitally skilled will start using it immediately.”* (P3, PM) Patients with minor complaints who want to avoid long wait times on the phone express enthusiasm about the app. Once patients create an account and use it once they remain using the app.

However, fewer patients than expected signed up for an account. A practice manager attributed this to the general resistance people have toward change: *“*T*hat’s because it’s simply a change. You know, people aren’t very fond of change. And I’m not saying my patient population is particularly conservative or progressive—it’s neither, really. Just people who don’t know it. They get the newsletter and say, ‘I think it’s a bit spammy; is the sender even legit? Hmm, maybe it’s fine, but it ended up in my spam folder.’”* (H1, GP)

Specific reasons for this hesitation could include patients’ preference for personal interaction: “*For example, patients who are vulnerable might come to the care chat app with a very simple question, but for them, it feels like a heavy burden, perhaps because of psychological problems.”* (H4, NP) Some patients may not feel the need for the app because their practice is already easily accessible. Moreover, certain patient groups might face greater challenges with an online application: *“So I think it’s fine for a large group of younger people up to around 40, but for those older than that, and especially for people from abroad who face a language barrier. We also have people with just low IQs—I think they’re not ready for this. And the elderly as well. I’d say it’s about 50/50.”* (H4, NP)

#### Changes in workflow

The impact of implementing Uw Zorg Chat on workflow was perceived as limited by most of the participants. One participant stated: *“What I mainly noticed was that patients were being scheduled during my consultation hours. Yes, I didn’t experience much inconvenience from it, just carried out my normal consultations.”* (H4, ANP)

However, other participants, particularly in practices that discontinued the pilot of Uw Zorg Chat, did see changes in workflow . These changes primarily involved adjusting schedules and supervision or correcting the work of Uw Zorg Chat. In the long term, DAs anticipated a more fundamental shift in their responsibilities. They predicted a significant reduction in telephone calls, which would require them to adopt new roles within the practice, such as independently running consultations. For GPs, it was expected that their consultations would predominantly consist of more complex cases, as simpler inquiries would already have been handled by the digital physician of Uw Zorg Chat.

The relative small changes following the implementation of Uw Zorg Chat also created an unawareness of the questions Uw Zorg Chat was managing: “*I also don’t think it’s used very much in our practice. You can see by the colors if it was from the chat. But I don’t really think it’s been used a lot. I haven’t really seen many colors in the agenda*.” (D3, DA). In the continuing practice this was made visible by sending weekly statistics: “*And then you see those statistics of how many people have been there, and then you think, ‘Oh yeah, that’s really nice indeed.’ But we haven’t really noticed that much*.” (D2, DA).

Uw Zorg Chat was promoted through various methods. Most promotional activities required only a one-time adjustment. However, actively promoting the chat service during phone calls proved to be a process of adaptation for some DAs. Not all DAs were immediately willing to actively promote this new approach. In some practices, personal motivation by practice managers was necessary to encourage them to adapt their methods: “*No, we didn’t do any advertising. There’s just no time for that. If you also have to explain all of that as well.”* (D5, DA)

The challenges in adapting to these changes might have been mitigated with a structured implementation plan. Two participants emphasized the importance of such a plan, particularly in retrospect, as no structured implementation plan had been established prior to the pilot: *“There should at least have been an implementation plan—one that you adjust every time to see if you can achieve success and to make agreements about when someone should be placed in the practice agenda and when not. What are the criteria, and how do you report back on things that aren’t going well? Now, this was just said to each other; people complained about it, but that doesn’t work. You have to say: this isn’t working; who is the project leader, and who will pick it up? Who is going to communicate it?”* (P3, PM).

### Effects

Many participants reported noticing limited effects and attributed this to the short duration of the pilot, they emphasized that the pilot needed more time than three months. They recommended a duration of at least one year to allow proper adoption. Also, the timing of the pilot was seen as unfortunate. Starting just before a holiday period led to reduced availability and made communication more challenging.

This outcome was different from what many participants had hoped or expected: *“No, you have to notice something in that time. Six weeks, I think, and then it was extended by another five or six weeks. But yeah, so that period of about three months—you should have been able to notice* something.” (H2, GP)

#### Positive Effects

The positive effects that were observed related mostly to reduced workload. This was mentioned by most of the participating practices: *“Yes, it was. It was much more manageable for us. We continued with it afterward, especially when the GP returned from vacation. Normally, Monday and Tuesday feel like a war zone, but this time, everything was handled beautifully. It went really well.”* (P3, PM)

Another positive effect was that the application allowed staff to take on other tasks, as a practice manager mentioned: *“Assistants mentioned: ‘it saves us time on making appointments and giving advice, so we can focus more on seeing our own patients.’ They’re fine with answering phones but not all day, so they also appreciate being able to do consultation hours and work with materials.”* (P3, PM)

The application also made the practice more robust: *“In that sense, we’re no longer as vulnerable to people who want more time off. I have a colleague who might want to go from four days to three. Then everything balances out again, and we can rely on the app as support, of course.”* (D1, DA)

The application also had a positive influence on the mentality of DAs: *“What I find great is that the assistants now think more creatively about how to organize work to make it easier. They’ve also learned to be stricter about calls between 8 and 10, except for making appointments. They now refer those calls to another time. If the emergency line rings and it’s not urgent, they say, ‘No, you’re on the emergency line. You need to call back on the regular line.’ They keep the emergency line free. That’s a win. They’ve learned that patients don’t mind being told to do things differently.”* (P3, PM)

Additionally, the patient population in consultations shifted, as frequent visitors could now ask their questions more easily via the app: *“I notice that a number of people who have the app now—I don’t see them as often. They handle everything through the app. They’re often people who get anxious quickly, have concerns, and can’t go to their grandmother to ask a question. Their grandmother sends them back, saying, ‘I’m not an expert, go to the doctor,’ and off they go. Perfect. It works very well.”* (H1, GP)

In the long term, one of the practice managers expected that the app might facilitate growth by reducing pressure: *“You don’t implement it to grow, but maybe at some point, it might encourage growth as a side effect.”* (P2, PM)

However, most of the effects were limited, with participants finding it difficult to draw firm conclusions: *“Otherwise, this part—has it saved me a lot of work? Well, I kept a small shadow record of that, and it wasn’t the case. Did it actually lead to significantly fewer phone calls? That’s also doubtful. Whether it did or not, I don’t know.”* (H3, GP)

#### Negative Effects

Negative effects primarily involved increased workload which was deemed frustrating by the practices that stopped the pilot: *“Then you look into it—hey, who did they have contact with? Can we retrieve that from the system? Were they billed properly? And then we found out that not everything had been billed correctly. It was just a bit of a waste because it also created extra work for us.”* (POH1, POH) And: *“They also called us sometimes. Or we had to call them back again, so we ended up doing double the work. It wasn’t like we could say, ‘Oh, how wonderful, we’re being relieved of work.’”* (D4, DA)

DAs as well as GPs mentioned that the reduction of workload was not necessarily satisfying. *“Now it’s nice to have some time for other things. You can catch up on overdue work. But eventually, it’s also nice to have full consultation hours again.”* (H4, GP)

Concerns were raised about accessibility of the GP: *“My parents would feel like they can’t reach their GP anymore. They’d just sit with their complaint. You have those kinds of people, too. I think we need to be cautious with that.”* (D2, DA)

Additionally, there were worries about providing incorrect information to patients: *“Patients would call or show up at the desk, waiting for an appointment for half an hour. They’d have an appointment through the app, but it wasn’t in the schedule, or it was scheduled for a week later.”* (POH1, POH)

All assistants in practices that did not continue the pilot felt that the changes to the schedule caused them to lose control: *“We were losing control because they started scheduling in emergency slots. Then, suddenly, someone would come in during the afternoon for eczema—something that could have been postponed. That happened regularly.”* (D5, DA)

### Wider context

#### Health insurance

The health insurer played a significant role in the implementation of this pilot. The insurer provided funding for the pilot and strongly supported its initiation: *“I know the program of our insurers is very much focused on addressing the GP shortage. They see a tremendous future for AI and the digitalization of GP practices as a way to tackle these shortages.”* (H3, GP) Another aim was to provide care for a large number of Leeuwarden residents unable to access a GP and to help practices grow.

Ultimately, a mismatch emerged between the expectations of the practices and those of the health insurer: *“They expected more than what was achieved. Someone from the insurer was present at the evaluation and said, ‘We’ve done our best to encourage you, but the results didn’t meet our expectations.’”* (P3, PM) That resulted in an end of funding the pilot: *“Unfortunately, the insurer said—something I wasn’t aware of—that the points raised during the interim evaluations were supposed to be addressed, but nothing was done. They said, ‘You had your chance, and we’re not continuing.’”* (P3, PM) GPs couldn’t afford it without help of the insurer, which resulted in an end of the pilot.

While the health insurer aimed to facilitate growth, none of the practices shared this goal. One significant reason was the impact of growth on evening, night, and weekend shifts, which increase with the number of patients: *“We’re not going to grow because GPs have to take on more shifts based on the size of their practice. That’s unmanageable. If you’re a solo practitioner, you already have full, demanding days, and then you also need to cover so many shifts—250 hours if you have 3,500 patients—on top of your regular work. That’s just not feasible.”* (P3, PM)

#### Cooperation participating practices

As mentioned before, three of the participating practices were part of a healthcare center. Poor collaboration between these practices hindered the effective exchange and implementation of feedback: *“People started with great enthusiasm, but you need structure when implementing changes. That structure was lacking in this healthcare center. A casual comment doesn’t lead to change or proper implementation. The wheel was reinvented multiple times across all practices. That wasn’t necessary. There should have been a central system in place, with fixed moments to address these issues.”* (P3, PM)

## Discussion

### Summary and comparison to existing literature

This study examined the implementation of a synchronous chat application in general practice and its impact on workload. The findings indicate that the application did not lead to a clear reduction in workload. While some participants reported a slight decrease in telephone traffic, they were hesitant to attribute this effect solely to the implementation of synchronous chat. Others experienced an increase in workload, primarily due to scheduling disruptions and the need to correct errors made by Uw Zorg Chat.

Notably, reports of increased workload and workflow disruptions were only observed in the three practices that discontinued the application, whereas the practice that continued its use did not report similar challenges. This suggests that organizational factors played a crucial role in the adoption and perceived effectiveness of the intervention.

Several organizational characteristics influenced the implementation’s success, including the degree of team engagement in the adoption process, the presence of structured feedback mechanisms and responsiveness to feedback, and the willingness to adapt existing workflows and integrate new processes. Collaboration with other practices within a healthcare center and the extent to which practices were willing to invest time and effort into the implementation process further influenced outcomes. Practice size was also identified as a key factor in adoption, with larger practices finding the implementation process easier.

Mostly concerns DAs rather than GPs or practice managers. Literature provided insight into these differing attitudes. A key facilitator in implementing digital innovations in healthcare is compatibility with existing systems (17, 18). Resistance is reduced when disruptions to daily workflows are minimized. One of the most significant workflow changes involved appointment scheduling, a task handled by DAs, but now partially managed by Uw Zorg Chat, which occasionally introduced scheduling errors. Since DAs were directly responsible for managing the agenda, they were most affected by these changes.

In much of the literature on facilitators and barriers to digital innovation, technical difficulties with new technologies are often cited as an obstacle. Successful adoption typically requires training for healthcare workers and a certain level of digital proficiency (18, 19). However, a key facilitator of this synchronous chat application is that it does not introduce these challenges. The tool can be implemented relatively easily without requiring adopters to develop new digital skills or become reliant on complex technology. Instead, the primary barriers lie in how the system integrates with existing workflows and interacts with other systems, rather than in the technical functionality of the application itself.

Another factor that contributed to the more negative perception among DAs compared to GPs was their level of involvement in the implementation process. Successful digital innovation adoption requires engaging the entire team and providing well set-up communication structures (17, 20, 21). In the discontinuing practices, this did not happen. The implementation was introduced rather abruptly, and opportunities for DAs to provide feedback or adjust to the new system were unclear, likely reinforcing their resistance. This could have also explained why DAs in the continuing practice did not report any challenges. Unlike in the discontinuing practices, they were actively involved in the decision-making process and received clear instructions on how to provide feedback. This proactive approach likely contributed to a smoother adaptation and greater acceptance of the new system.

Another explanation for the difference in attitude among DAs in the continuing and discontinuing practices could have been their perceived added value of synchronous chat in the general practice. DAs in the discontinuing practices stated that they did not see any benefit in using the application, whereas those in the continuing practice did. Willingness to change often depends on recognizing the need for change (17, 18, 21, 22). The DAs in the discontinuing practices did not see a need for change, which may have contributed to their resistance.

While GPs and practice managers acknowledged some reduction in workload and expressed interest in further exploring the potential of Uw Zorg Chat, financial constraints ultimately hindered continuation. The withdrawal of financial support from the health insurer made the costs unsustainable, leading to the decision to discontinue its use. Systematic reviews on factors influencing the implementation of electronic health technologies consistently highlight cost as one of the most significant barriers (17, 19). The practice that continued using the app had twice as many patients as those that discontinued, suggesting that larger practices may have more resources to support new innovations. The practices that discontinued the pilot explicitly identified their smaller size as a limiting factor in adopting new technologies. The initial investment was perceived as substantial, while the return on investment remained uncertain and gradual due to limited patient adoption. For smaller practices, this financial pressure was especially difficult to manage, as they had fewer financial buffers to cover expenses and sustain operations until the app became financially viable. This pattern is also reflected in broader research on EHR implementation, where cost is frequently cited as a primary barrier, particularly for smaller practices (23).

Financial support is an important incentive in enabling (smaller) practices to adopt new technologies (17, 18, 23). This was also evident in this study, where initial funding by the health insurer encouraged participation in the pilot. However, when financial support was withdrawn, practices were unable or unwilling to continue using the app, highlighting the dependency on external funding. Literature identified several facilitators to financial support, one of which is the level of commitment of the project leader (24). Funders sought dedication and conviction of project leaders before committing to long-term investments. In the case of Uw Zorg Chat, GPs showed limited willingness to adapt their workflows, such as closing phone lines or actively participating in evaluations, which led to the insurer’s decision to withdraw funding, ultimately resulting in the termination of the pilot.

There are several explanations from the literature for the limited willingness to adapt workflows. Leaders are more likely to provide support when they are actively involved and regularly updated on the project’s progress (18-20). In the practice that continued using Uw Zorg Chat, weekly updates on patient registrations and message activity helped maintain enthusiasm by providing clear insights into the app’s impact. In contrast, the lack of such communication in the discontinuing practices may have led to decreased motivation, as it was unclear how much work Uw Zorg Chat was actually taking over. Moreover, in two of the four participating practices, the person responsible for supervising the DAs was not engaged throughout the process. Given that leadership commitment is a key facilitator for successful implementation, this lack of involvement may have hindered adoption.

Another barrier to adopting remote care is the unwillingness of healthcare workers to lose personal connection (21). GPs who were reluctant to adjust their workflows cited this as the primary reason for refusing to close phone lines, emphasizing the value of direct, personal interaction with their patients.

Besides variations in the success of implementation, a key consideration when introducing synchronous chat was the burden of ENW shifts. Discontinuing practices cited this, alongside cost, as a primary barrier to implementation. While the health insurer viewed growth as a key objective, practices found this expectation unrealistic. Therefore, it is essential to align goals beforehand and assess their feasibility. If growth was a primary objective, the implications of ENW shifts had to be explicitly addressed as part of the discussion.

It remains difficult to draw definitive conclusions on effectiveness, as the limited number of patients who created an account and used Uw Zorg Chat resulted in minimal observable impact.

### Implications for research and practice

The phenomenon of pilotitis, where digital health innovations remain stuck in the pilot phase without scaling up, is a persistent challenge in healthcare. To avoid this, implementation efforts must shift from short-term testing to long-term sustainability. One of the key factors in achieving this is providing financial information (25). As seen in this study, practices that discontinued the use of Uw Zorg Chat struggled with costs once initial funding was withdrawn. Future implementations should take in consideration the financial situation of general practices on the long-term, even when the supplier withdraws funding. Additionally, larger practices had an advantage in absorbing costs, suggesting that financial models should account for differences in practice size.

For practices looking to implement digital health technologies, this study highlighted that compatibility was not just a technical issue but an organizational one. Simply ensuring that a new tool integrates with existing systems was not enough; structured feedback mechanisms and leadership engagement were essential for managing workflow disruptions effectively. Clear implementation plans with defined goals and evaluation strategies should be in place. Practices should provide communication channels where staff can voice concerns and contribute to solutions, particularly for DAs. GPs and practice managers must take an active role in guiding the adaptation process by providing regular updates and addressing challenges transparently.

To improve adoption, it is important to ensure communication structures with the provider and adopter as well as within the system of the adopter. Additionally, gaining a deeper understanding of existing workflows, such as through structured meetings with general practices and representatives of the application, could help align the system more effectively with daily operations.

Future research should explore why patients faced difficulties in registering for the app. While this study provides initial insights, it is essential to verify these findings directly with patients. Additionally, assumptions made by healthcare providers, such as patients disliking remote care or older adults being reluctant to use chat applications, should be examined to determine whether they reflect actual patient experiences.

Because only a limited number of patients created an account, effect measurement should be repeated after more time has passed to assess its long-term effects.

### Strengths and limitations

This study had several strengths and limitations worth noting. One strength was that the majority of interviews were conducted face-to-face in Leeuwarden. This in-person setting created a sense of trust and connection between the interviewer and participants. Participants indicated that they appreciated the opportunity to interact with someone in person, which encouraged openness in sharing both positive and negative thoughts.

Additionally, the study included a diverse participant population. It involved individuals of various ages, with both short-term and long-term employment histories, and represented practices that continued as well as those that discontinued the use of Uw Zorg Chat. This diversity enhanced the transportability of findings by capturing a wide range of experiences and perspectives.

Another strength of this study was that it examined the entire implementation process instead of just small parts of it. Because so many aspects were considered, the findings provided a full picture rather than scattered insights. This strengthened the conclusions and made the recommendations more reliable for future implementations of synchronous chat consultations.

However, the study also had several limitations. One notable limitation was the potential for response bias due to the perceived relationship between the interviewer and Uw Zorg Chat. Participants were aware that the interviewer had previously worked for Uw Zorg Chat and viewed her as a representative of the organization. This association could have made it more challenging for participants to provide critical feedback, as they might have been hesitant to express negative experiences out of concern for appearing personal.

To mitigate this, participants were encouraged to share both positive and negative experiences openly, and interviews were structured to create a neutral and nonjudgmental setting. However, this association with Uw Zorg Chat may have also explained the reluctance of some practices that had discontinued using the application to participate in the study. It is possible that those practices had negative experiences with Uw Zorg Chat and found it uncomfortable to engage with someone affiliated with the organization. As a result, valuable insights may have been missed. However, none of the participants in this study mentioned this association as a barrier to speaking freely.

Additionally, another limitation of this study was the lack of interviews with both patients and employees of Uw Zorg Chat. While healthcare providers offered explanations for why patient registrations for the application were limited, these explanations should have been confirmed by direct input from patients themselves. Furthermore, understanding the well-being and experiences of Uw Zorg Chat employees would have been critical in determining whether the application was sustainable for future implementation.

## Conclusion

This study examined the implementation of a synchronous chat consultation system in general practice, highlighting both facilitators and barriers to adoption. While the application had the potential to reduce workload by managing simple inquiries remotely, its success depended largely on organizational factors. Strong leadership commitment, compatibility with existing systems and feedback structures facilitated adoption. Financial constraints were considered the main barrier, as smaller practices struggled to sustain costs once initial funding ended. Other barriers included workflow disruptions, administrative burden, and limited involvement of doctor’s assistants. Although the limited number of patient registrations made it difficult to assess the full impact of the tool, early indications suggested that a synchronous chat consultation system can help alleviate staff workload if close attention is paid to organizational structures and the implementation plan.

## Supporting information

Supplementary file 1. Interview guide

Supplementary file 2. Interview excerpts

## Data Availability

All data produced in the present study are available upon reasonable request to the authors through DataverseNL.

https://doi.org/10.34894/FNZUDY

