## Supplementary file 1. Interview guide for "Implementation and evaluation of synchronous chat in general practice - a qualitative interview study"

**Interview Guide CHAT-GP**
Version 2.0, 11-12-2024

This interview guide describes a qualitative research study using semi-structured interviews for the study:
*"Implementation and evaluation of chat consultations in general practice: a qualitative interview study."*

The interviews will last approximately 45-60 minutes.

Participants are approached with a request to take part in a face-to-face interview based on their experiences working in one of the general practices involved in a pilot project (*Uw Zorg Chat*), which provided patients with an online option to chat with a healthcare provider from an external practice.

**Objective**

The aim of these interviews is to gain insight into personal experiences with the implementation process of *Uw Zorg Chat*, the observed effects on work, and expectations for the future.

**Preparation**

Before starting the interview, discuss:

- Introduction of the interviewer and explanation of the study/research objective
- Emphasizing that participation is voluntary and that the participant may stop at any time
- Asking for consent to record the interview

Start the recording in Teams and state the date and participant number.

*"We will process and analyze the answers anonymously so that they cannot be traced back to you. Therefore, we are recording the interview, but this will also be done anonymously. There are no right or wrong answers in this interview. You may indicate at any time if you wish to stop."*

**Questions**

**General Information**

- Age, gender
- Role in the general practice, years of experience in this role, and time working in the practice that participated in the pilot
- Experience with digital healthcare: e-consultations, email, telephone, video calls, digital triage, telemonitoring?

**Implementation Process**

- Why did you want to participate in this pilot?
- What were your expectations of the pilot?
- What did you think about the decision to conduct the pilot?
- How did the team react to working with this application?
- How were you/the team involved in starting the pilot?
- How did patients react to the pilot?
  - How were patients directed to the app?
- In what way was the app intended to be used?
  - Is it being used in that way?
- How does it integrate with the work?
  - How is it used/when/by whom?
  - What oversight is in place?
  - How have work processes changed due to the app's use?
  - What do you notice about its use in daily practice?
- What do you think of the integration of the app in your general practice?
- How were you supported in implementing the app? How was that experience?
- What are the key lessons you learned from this implementation process?

**Effects**

- What is your overall impression?
- Have you noticed changes in your work due to the pilot?
- What are the positive effects?
  - How has it affected workload?
- What are the negative effects?
  - Do you find it safe?
  - Does it have a pull effect (i.e., attract more demand)?
  - Does the app generate more work? If so/not, why?
  - How did you feel about someone else providing care for your patients?
  - Has the nature of cases in your consultation hours changed?
- What impact does it have on continuity of care?
- Does it achieve the goal for which it was implemented? Why?

**Future**

- Would you recommend this app? Yes/no? If yes, to whom?
- How do you see the future of this app?
- What are your recommendations?

Do you have any additional comments?

**Conclusion**

*"Thank you very much for your cooperation."*
